# Neutralizing responses in fully vaccinated with BNT162b2, CoronaVac, ChAdOx1, and Ad26.COV2.S against SARS-CoV-2 lineages in Colombia, 2020-2021

**DOI:** 10.1101/2022.03.15.22272371

**Authors:** Diego A. Álvarez-Díaz, Ana Luisa Muñoz, María T. Herrera-Sepúlveda, Pilar Tavera-Rodríguez, Katherine Laiton-Donato, Carlos Franco-Muñoz, Héctor Alejandro Ruiz-Moreno, Dioselina Pelaez-Carvajal, Alejandra M. Muñoz-Suarez, Marisol Galindo, Jhonnatan Reales-Gonzalez, Jenssy D. Catama, Beatriz Helena De Arco, Tatiana Cobos, Edgar J. Arias-Ramirez, Marcela Mercado-Reyes

## Abstract

**Background:** By March 2022, around 34 million people in Colombia had received a complete scheme of vaccines against the severe acute respiratory syndrome coronavirus 2 (SARS-CoV-2) including, mRNA-based vaccines, viral vectored coronavirus vaccines, or the inactivated whole virus vaccine. However, as several SARS-CoV-2 variants of concern (VOC) and interest (VOI) co-circulate in the country, determining the resistance level to vaccine-elicited neutralizing antibodies (nAbs) is useful to improve the efficacy of COVID-19 vaccination programs.

**Methods:** Microneutralization assays with the most prevalent SARS-CoV-2 lineages in Colombia during 2020-2021 were performed using serum samples from immunologically naïve individuals between 9 and 13 weeks after receiving complete regimens of CoronaVac, BNT162b2, ChAdOx1, or Ad26.COV2.S. The mean neutralization titer (MN50) was calculated by the Reed–Muench method and used to determine differences in vaccine-elicited nAbs against the SARS-CoV-2 lineages B.1.111, P.1 (Gamma), B.1.621 (Mu), and AY.25.1 (Delta).

**Results:** The most administered vaccines in the country, BNT162b2 and CoronaVac, elicited significantly different nAb responses against Mu, as the GMTs were 75.7 and 5.9-fold lower relative to the control lineage (B.1.111), while for Delta were 15.8 and 1.1-fold lower, respectively. In contrast, nAb responses against Mu and Delta were comparable between ChAd0×1-s and Ad26.COV2.S as the GMTs remained around 5 to 7-fold lower relative to B.1.111.

**Conclusions:** The emergence of SARS-CoV-2 variants in Colombia with a significant capacity to escape from vaccine-elicited nAbs indicates that a booster dose is highly recommended. Furthermore, other non-pharmacological measures should be retained in the vaccinated population.

## Introduction

Five vaccines against the severe acute respiratory syndrome coronavirus 2 (SARS-CoV-2) are authorized in Colombia; by the second week of March 2022, nearly 79 million vaccine doses had been administered. The inactivated whole virus vaccine CoronaVac (Sinovac Life Sciences, Beijing, China) and the BNT162b2 (Pfizer-BioNTech), an mRNA-based vaccine that encodes the SARS-CoV-2 full-length spike (S) gene, are the two main vaccines administered in the country representing 31.5% and 30.3% of the doses, respectively. In addition, the ChAdOx1 (Oxford-AstraZeneca) and Ad26.COV2.S-Janssen (Johnson &Johnson) vaccines which are based on replication-incompetent adenoviral vectors, expressing a variant of the SARS-CoV-2 S protein account for 15.5% and 8.5% of the administered doses, respectively. Finally, the mRNA-1273 vaccine (Moderna), another mRNA-based vaccine that encodes the SARS-CoV-2 full-length S gene of SARS-CoV-2, accounts for 14.2% of the doses administered in the country.

However, around 34 million people have received the complete scheme of vaccination which is two doses for CoronaVac, BNT162b2, ChAdOx1 or Moderna, and one dose for Ad26.COV2.S, while only 9.3 million have received a booster dose with a homologous or heterologous vaccine (1, 2).

The routine genomic surveillance of SARS-CoV-2 in Colombia allowed the identification of variants of interest (VOI) and concern (VOC) that co-circulate in the country with evidence of scape to neutralizing antibodies (nAb) generated by the BNT162b2 vaccine or natural infection (3, 4). The first two COVID-19 epidemic peaks in Colombia were dominated by B.1 non-VOC/VOI lineages (B.1.111, B.1.420, and B.1) and occurred from June to September 2020, and December 2020 to January 2021, respectively, while between April to August 2021, the third epidemic peak was dominated by Gamma (P.1) and Mu (B.1.621), representing 25% and 49% of the total cases (3). These two variants were gradually displaced by Delta lineages from July to December 2021. Soon afterward, the arrival of Omicron before the end of December 2021, during the fourth epidemic peak, resulted in the total displacement of Delta lineages by the end of January 2022 (5).

The Gamma variant was the first VOC detected in the country. It has 12 mutations in the S protein, including critical substitutions on the Spike RBD (K417N, E484K, and N501Y) associated with an increment in the binding affinity to the human ACE2-receptor (6) and reduced levels of neutralizing antibodies (3, 6-8). Meanwhile, the Mu variant contains a distinctive profile of mutations in the S protein (D614G, D950N, E484K, ins145N, N501Y, P681H, R346K, T95I, Y144T, Y145S) (9), involved in the resistance to vaccine-elicited or natural infection-elicited antibodies (3, 10). Finally, almost one hundred Delta variant (B.1.617) sublineages were detected in Colombia, five of them (AY.20, AY.25, AY.26, AY.3, AY.5) with higher frequency. The dominant sublineage, AY.25, also carries multiple mutations at the S protein (D614G, del157/158, D950N, L452R, P681R, T19R, T478K, V1065L) associated with immune evasion (11-13).

Systematic reviews and meta-analyses of phase III studies on vaccine effectiveness before the emergence of variants of concern suggest respectively an efficacy against symptomatic and severe COVID-19 of 82%-100% and 75%-95% for BNT162b2, 98%-100% for mRNA-1273, 66%-81%, and 100% for ChAdOx1, 72%-74% and 85% for Ad26.COV2.S and, 50%-83% and 100% for CoronaVac (2). However, the emergence of SARS-CoV-2 VOCs and VOIs carrying genetic markers at the S protein associated with greater transmissibility rate and resistance to nAbs at the end of December 2020, challenged this scenario. Several studies around the world evidenced a substantial fold decrease in the geometric mean nAb titer (GMT) against different VOCs/VOIs, relative to ancestral SARS-CoV-2 lineages in individuals with complete regimens of the vaccines currently approved or authorized in Colombia (2). Furthermore, studies on nAb responses in BNT162b2-vaccinated in Colombia evidenced a robust reduction in the neutralization of Mu, a VOI first detected in the Caribbean region of the country (9), by 75.7- and 17.7-fold, relative to B.1.111 and Gamma, respectively, which was comparable to reports from Asia and Europe (3). As these reductions in nAbs responses ranged from minimal up to total escape from vaccine nAbs depending on the SARS-CoV-2 variant and the population evaluated (2, 14), it is important to characterize the profile of vaccine nAbs in the context of the circulating SARS-CoV-2 variants in each country. Hence, we used a microneutralization assay with infectious viruses to determine the nAb responses against B.1.111, Gamma, Mu, and Delta in individuals fully vaccinated with four out of the five vaccines authorized in Colombia, altogether accounting for 86% of the doses administered: BNT162b2, CoronaVac, ChAdOx1 and Ad26.COV2.S.

## Methods

### Patients and samples

Serum and nasopharyngeal swab samples were collected from individuals with written informed consent approved by the Ethics Committee of the Colombian National Health Institute (CEMIN)-04-2021. Serum samples were collected from immunologically naïve individuals between 9 and 13 weeks after receiving the complete regimens of BNT162b2 (n = 31; 3 males and 28 females, age range, 23–62 years), CoronaVac (n = 30; 9 males and 21 females; age range 18–58 years), ChAdOx1 (n = 20; 10 males and 10 females; age range 50 –81 years) or Ad26.COV2.S (n = 26; 9 males and 17; age range 21 –61 years).

### Virus isolation

Nasopharyngeal swab samples with positive RT-PCR for SARS-CoV-2, complete genome sequencing and PANGO lineage assignment B.1.111 (EPI_ISL_526971), P.1/Gamma (EPI_ISL_2500971), B.1.621/Mu (EPI_ISL_1821065) and AY.25.1/Delta (EPI_ISL_7314401), were selected for virus isolation and used for inoculation of Vero E6 monolayers as described previously (3). All assays with infectious SARS-CoV-2 viruses were performed in a biocontainment laboratory level 3.

### Microneutralization and binding antibody assays

Microneutralization assays with infectious viruses were performed in Vero E6 monolayers as described (4), by incubating 120 mean tissue culture infectious doses (TCID50) of SARS-CoV-2 isolates belonging to the lineages B.1.111, P.1 (Gamma), B.1.621 (Mu), and AY.25.1 (Delta), with two-fold serial dilutions (1:4 to 1:2560) of sera from vaccinated volunteers. Cytopathic effect was examined after five days of incubation and the mean neutralization titer (MN50) was calculated by the Reed–Muench method (15). The absence of total (IgG/IgM) anti-S antibodies before vaccination was verified using the SARS-CoV-2 Total assay (COV2T, Siemens Healthcare Diagnostics Inc., NY, USA). The absence of IgG anti-nucleoprotein antibodies during clinical follow-up of individual vaccinated with BNT162b2, ChAdOx1, Ad26.COV2.S was verified using the qualitative ELISA ID Screen SARS-CoV-2-N IgG Indirect (ID Vet, Montpellier, France), following the manufacturer’s instructions. Subsequently, the concentration of anti-spike IgG antibodies was tested using the SARS-CoV-2 IgG assay (sCOVG, Siemens Healthcare Diagnostics Inc., NY, USA) on the ADVIA Centaur XPT platform (Siemens). The cut-off value was defined as reactive ≥ 1.0 U/mL. Index values were expressed in binding antibody units per milliliter (BAU)/mL using the conversion factor (WHO standard) of 21.8, as determined for the Siemens assays (16).

### Statistical analysis

Differences between the mean neutralization titer (MN50) for BNT162b2, CoronaVac, ChAdOx1, and Ad26.COV2.S against each SARS-CoV-2 variant were determined using the Kruskal–Wallis test, followed by Dunn’s post hoc test for multiple comparisons where a p-value of <0.05 was statistically significant. An arbitrary value of 2 was assigned to samples with MN50 <4. Statistical analysis was performed using GraphPad PRISM 6.0.1 (GraphPad Software, San Diego CA, USA).

## Results

### Vaccine elicited Anti-S IgG antibody titers correlate with neutralizing antibody titers

No significant differences were observed between the binding anti-S IgG antibody titers elicited by CoronaVac, ChAdOx1, and Ad26.COV2.S. However, the titers elicited by BNT162b2 were significantly higher than the other three vaccines (Fig. 1). Neutralizing antibody titers elicited by BNT162b2, CoronaVac, ChAdOx1, and Ad26.COV2.S against B.1.111, Mu, and P.1, correlated well with total binding anti-S IgG antibody titers. For the Delta variant, all vaccines with exception of BNT162b2 showed a correlation of nAb and total binding antibodies (Table 1).

**Table 1.** Comparison of neutralizing and binding Anti-S IgG antibody titers

| Vaccine | GMBA titers<br>(BAU/mL)/(95%<br>CI) | Comparison |  | Spearman r* | p value |
| --- | --- | --- | --- | --- | --- |
| BNT16b2 | 1353<br>(959.9 – 1907) | MN50 B.1.111 | Anti-S IgG titer BAU/mL | 0.713 | < 0.0001 |
|  |  | MN50 Gamma (P.1) | Anti-S IgG titer BAU/mL | 0.503 | 0.0039 |
|  |  | MN50 Mu (B.1.621) | Anti-S IgG titer BAU/mL | 0.690 | < 0.0001 |
|  |  | MN50 Delta (AY.25) | Anti-S IgG titer BAU/mL | 0.255 | 0.1655 |
| CoronaVac | 111.3<br>(76.31 – 162.4) | MN50 B.1.111 | Anti-S IgG titer BAU/mL | 0.709 | < 0.0001 |
|  |  | MN50 Gamma (P.1) | Anti-S IgG titer BAU/mL | 0.453 | 0.0118 |
|  |  | MN50 Mu (B.1.621) | Anti-S IgG titer BAU/mL | 0.650 | < 0.0001 |
|  |  | MN50 Delta (AY.25) | Anti-S IgG titer BAU/mL | 0.563 | 0.0012 |
| ChAdOx1-s | 348.6<br>(174.8 – 695.3) | MN50 B.1.111 | Anti-S IgG titer BAU/mL | 0.496 | 0.0259 |
|  |  | MN50 Gamma (P.1) | Anti-S IgG titer BAU/mL | 0.761 | < 0.0001 |
|  |  | MN50 Mu (B.1.621) | Anti-S IgG titer BAU/mL | 0.811 | < 0.0001 |
|  |  | MN50 Delta (AY.25) | Anti-S IgG titer BAU/mL | 0.826 | < 0.0001 |
| Ad26.COV2.S | 78.36<br>(55.2 – 115.4) | MN50 B.1.111 | Anti-S IgG titer BAU/mL | 0.625 | 0.0008 |
|  |  | MN50 Gamma (P.1) | Anti-S IgG titer BAU/mL | 0.725 | < 0.0001 |
|  |  | MN50 Mu (B.1.621) | Anti-S IgG titer BAU/mL | 0.589 | 0.0019 |
|  |  | MN50 Delta (AY.25) | Anti-S IgG titer BAU/mL | 0.779 | < 0.0001 |

GMBA: geometric mean titer of binding antibodies. \*Two-tailed

**Figure 1.**
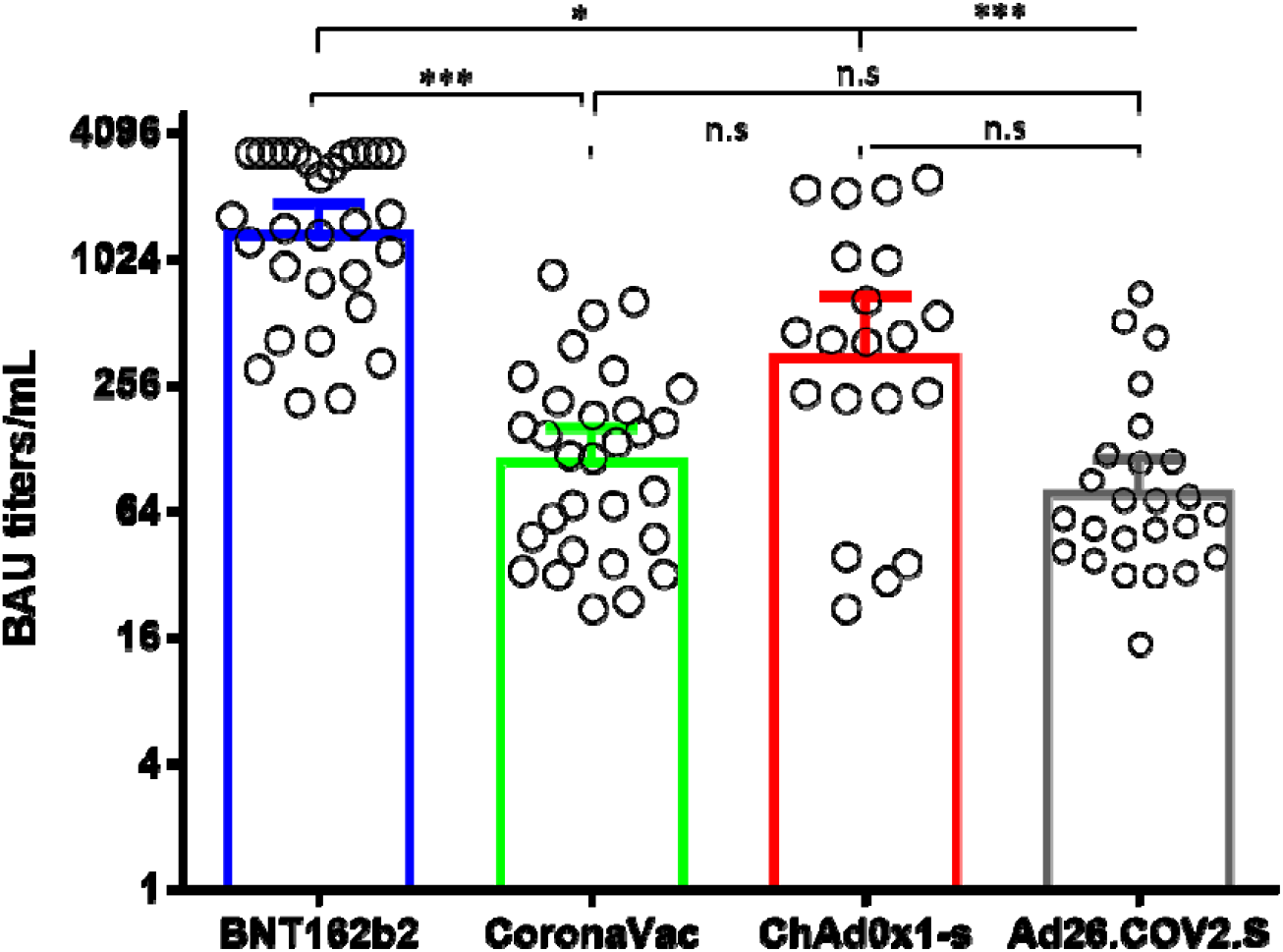
Comparison of binding Anti-S IgG antibody titers. BAU: binding antibody units. *: p = 0.01, ***: p < 0.0001. n.s: no significant

### Reduced nAb responses against Mu and Delta variants in vaccinated individuals with BNT162b2, CoronaVac, ChAdOx1, and Ad26.COV2.S

Microneutralization assays with sera from vaccinated individuals with BNT162b2, ChAdOx1, and Ad26.COV2.S revealed an overall reduction of the GMT against B.1.621 (Mu) and AY.25.1 (Delta), relative to B.1.111 and P.1 (Gamma) (Fig. 2a). In contrast, the GMT for CoronaVac was significantly lower only for Mu (Fig. 2c). Thus, while the overall GMT for B.1.111 and P.1 (Gamma) ranged between 51.7 and 401.3, for B.1.621 (Mu) and AY.25.1 (Delta) ranged between 5.3 and 46.6 (Table 2). This reduction was more pronounced against the B.1.621 (Mu), in individuals vaccinated with BNT162b2 and CoronaVac, as the GMT and seropositivity rate were lower among all the vaccines and variants tested (Table 2). Neutralizing antibody titers from all the tested vaccines were uniformly distributed against AY.25.1 (Delta) because when compared, only slight significant differences were observed in the GMTs between CoronaVac and Ad26.COV2.S (Fig. 2a). The highest nAb response was induced by BNT162b2 against B.1.111 (Fig. 2b), followed by ChAdOx1 which showed GMT titers above 200 against B.1.111 and P.1 (Gamma) (Fig. 2d). Furthermore, ChAdOx1 showed a higher GMT and the best performance against B.1.621 (Mu) as in contrast to the other vaccines, the seropositivity rate against this variant was 100% (Fig. 2a and 2d).

**Table 2.**
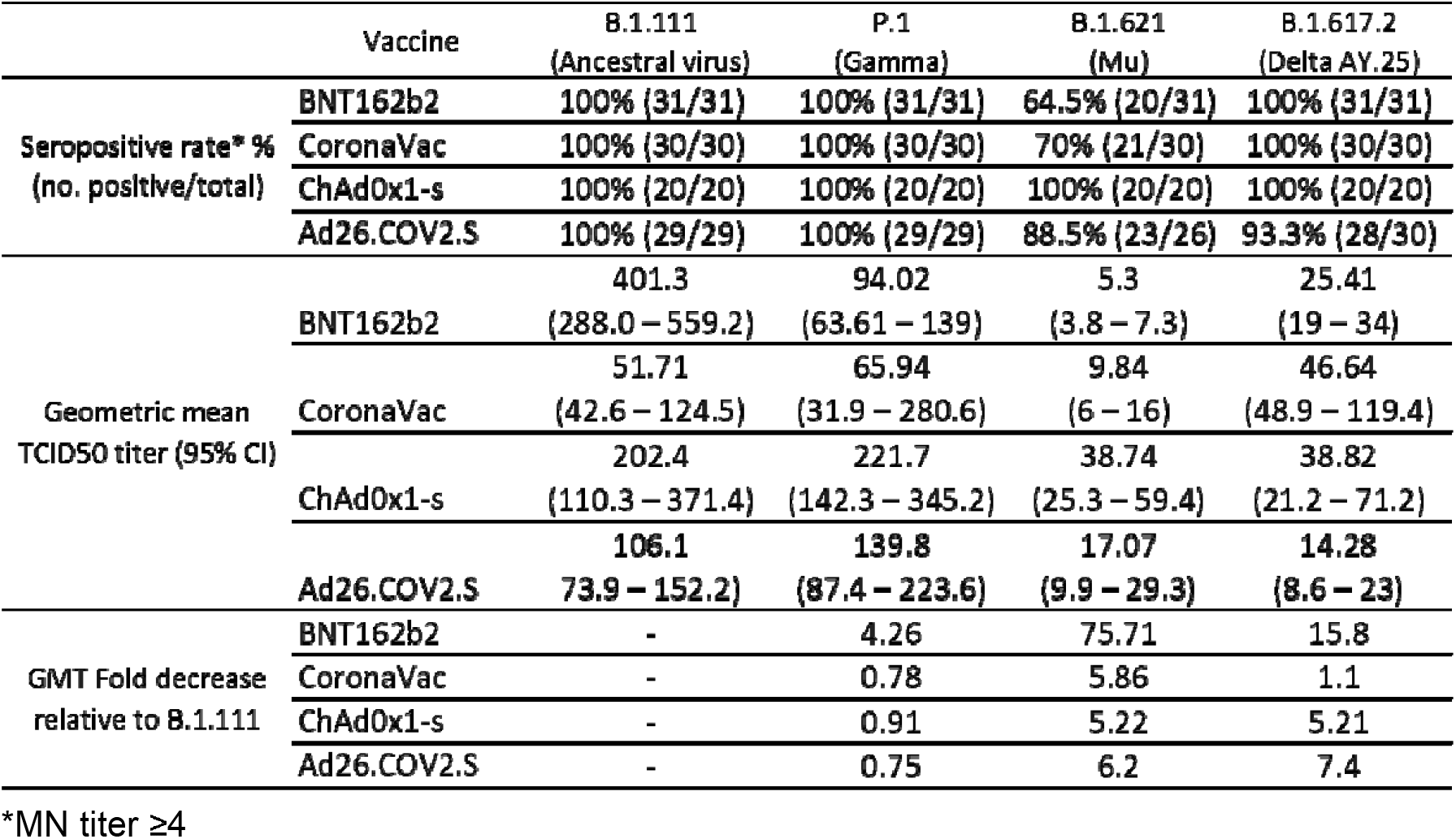
Summary of vaccine antibody responses against SARS-CoV-2 variants

**Figure 2.**
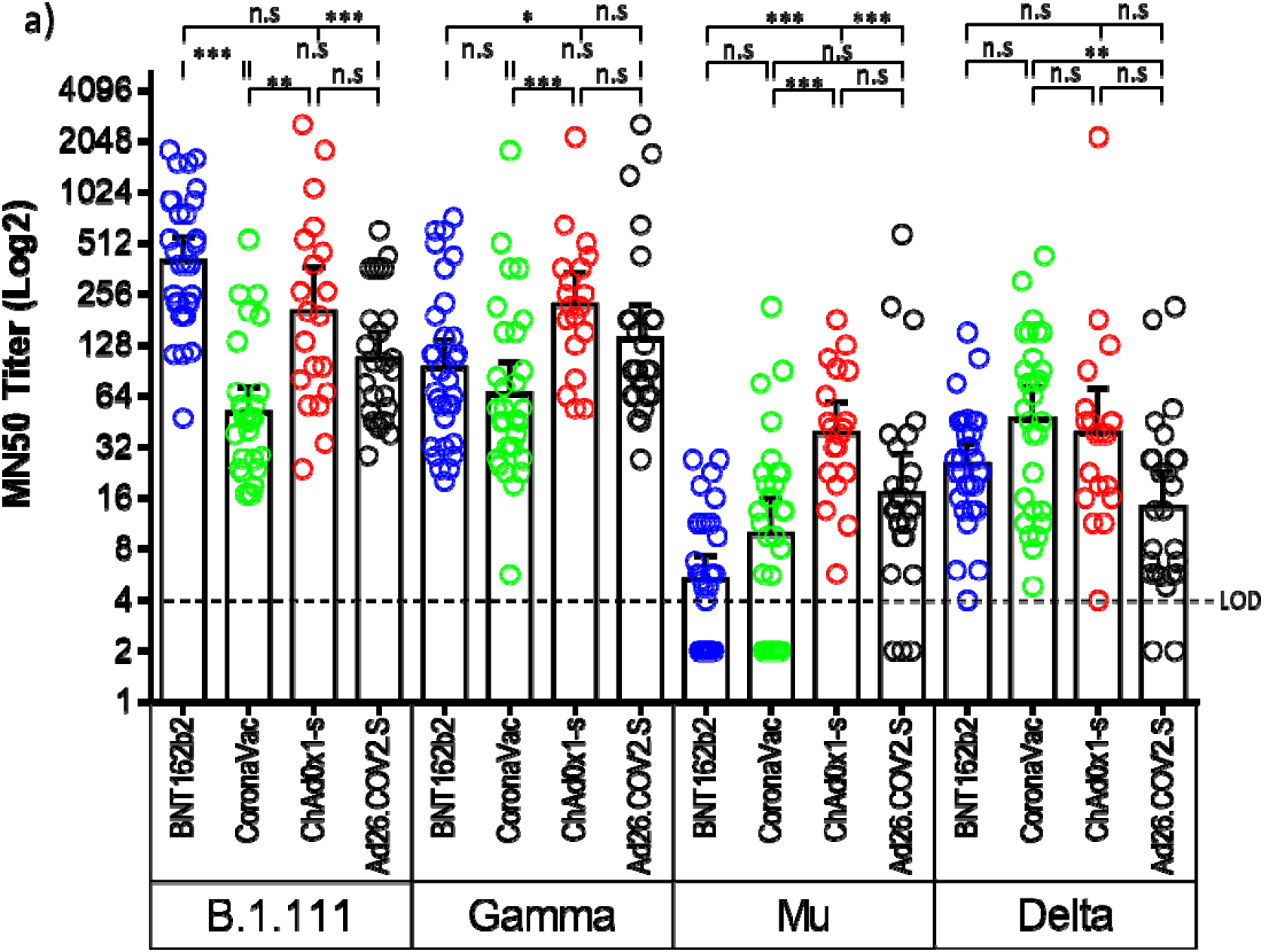
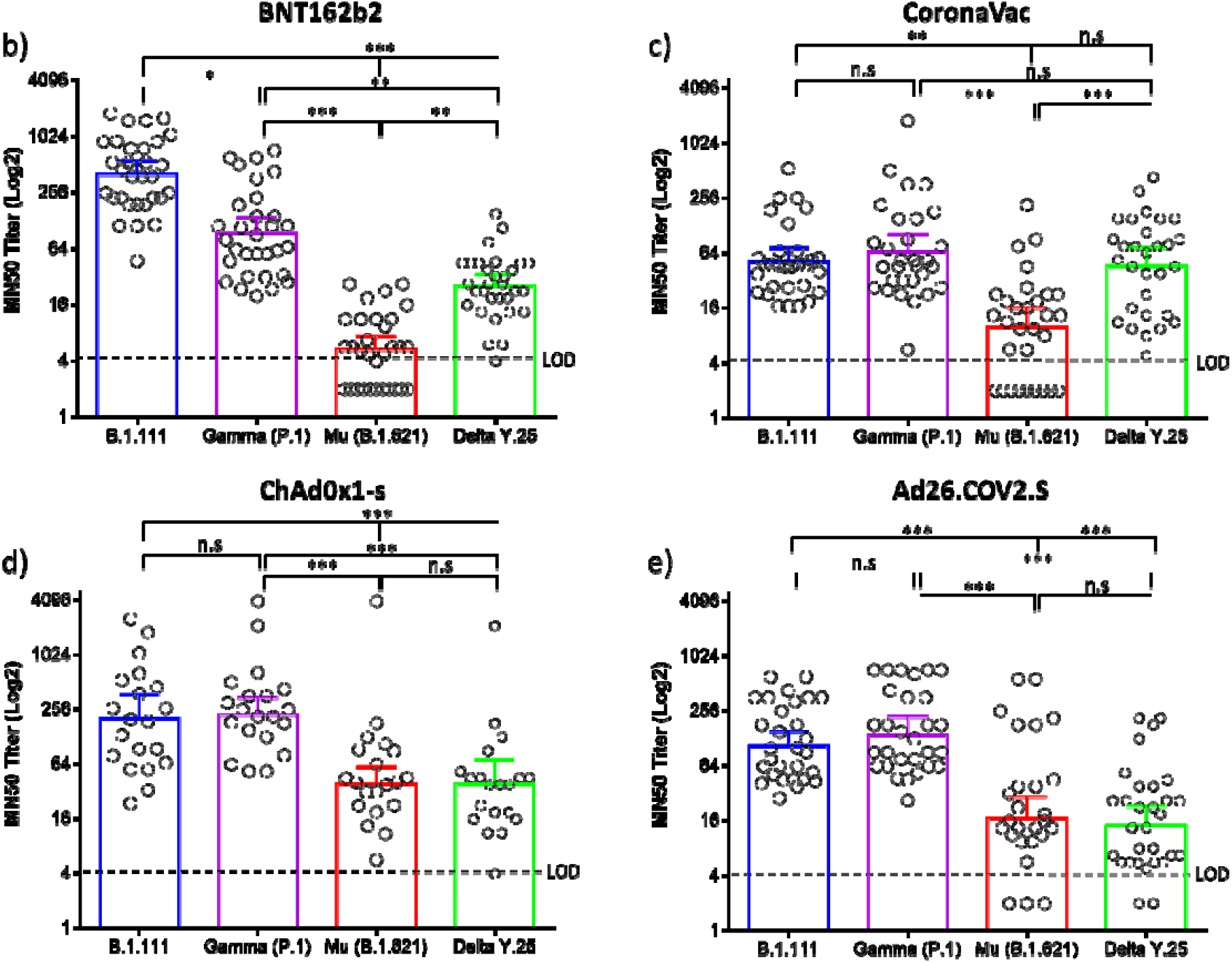
Comparison of neutralizing activity in BNT162b2, CoronaVac, ChAdOx1, and Ad26.COV2.S -vaccinated individuals by SARS-CoV-2 variants. a) Comparison of GMTs between vaccines against each SARS-CoV-2 variant. b, c, d, and e) Comparison of GMTs between SARS-CoV-2 variants for each vaccine. *: p = 0.01, **: p < 0.001, ***: p < 0.0001. n.s: no significant. LOD: limit of detection.

## Discussion

Massive vaccinations against SARS-CoV-2 around the world began in December 2020, reducing the rate of COVID-19-related hospitalizations and deaths (17, 18). However, the emergence of VOC and VOI challenged the effectiveness of available COVID-19 vaccines, and surveillance of their performance became a priority. This surveillance should include studies across different populations to estimate the global effectiveness of SARS-CoV-2 vaccines (19), as well the reconsideration of vaccination programs and the redesign of existing vaccines.

In general, reduced nAb responses were observed against Gamma, Mu, and Delta depending on the vaccine tested. However, nAb responses elicited by each vaccine were significantly reduced against the VOI Mu relative to B.1.111 and Gamma, which could imply an overall reduced efficacy of the vaccines BNT162b2, ChAdOx1, and Ad26.COV2.S, to prevent the infection with this variant. Studies in individuals vaccinated with BNT162b2 evidenced reduced nAb responses against pseudoviruses harboring the spike protein of the Mu variant, relative to four VOCs (Alpha, Beta, Gamma, and Delta) and two VOI (Epsilon and Lambda) (10). Furthermore, studies with infectious viruses also evidenced reduced nAb responses in BNT162b2 vaccinated patients (3).

Genetic, epidemiologic, and host factors can contribute to the notorious ability of the Mu variant to escape from nAbs, however, a profile of mutation of interest shared with Omicron (T95I, R346K, N501Y, P681H) could explain this phenomenon as no nAb responses were observed against Omicron with samples from individuals vaccinated with BNT162b2 or CoronaVac (14). In line with this, studies with pseudoviruses evidenced that neutralization resistance of the Mu variant to nAbs elicited by Ad26.COV2.S, mRNA-1273, and BNT162b2 was caused by R346K and E484K mutations (20).

Furthermore, in this study was evidenced a reduction of BNT162b2 elicited nAb titers against Gamma by 4.26-fold, while no significant differences were observed for CoronaVac, ChAdOx1, and Ad26.COV2.S relative to the B.1.111 lineage. In contrast, Dejnirattisai W., et al evidenced a reduction of nAb titers by 2.6 and 2.9-fold for BNT162b2 and ChAd0×1-s against Gamma relative to a control strain (7). These differences could be explained by differences in the virus strain as well as sampling times, as Dejnirattisai W., et al evaluated nAb responses 7–28 days following the second dose, while in the present study, nAb responses were evaluated 60–90.

As for the Delta variant, our results were comparable to studies performed in Asia, Europe, and North America, which evidenced resistance of the Delta variant to nAbs after receiving two doses of BNT162b2 and ChAd0×1-s (10-13).

Despite multiple studies evidence differential vaccine-elicited nAb responses against SARS-CoV-2 variants, the impact of nAb titer on the clinical outcome is not fully understood. It is known that higher nAb levels have a positive correlation with protection against COVID-19 (21). However, remains to be resolved the protective titer of neutralizing antibodies, Lau, E.H.Y., et al reported that a 1:40 hemagglutination-inhibition antibody titer protects from infection against influenza (22). Moreover, Gharbharan A. et al., suggest that a titer of 1:80 or higher after the convalescent plasma transfusion for the treatment of COVID-19 inhibits the viral growth *in vitro* by 95% (23). For these reasons, we hypothesize that a titer of 1:80 or higher may protect against progression to severe disease, which, for public health reasons, should be achieved preferably using vaccination.

These results highlight the importance of serological tracking of infected or vaccinated people for public health decision-making, along with genomic surveillance programs to identify the introduction of new SARS-CoV-2 variants. Furthermore, SARS-CoV-2 vaccines may be updated periodically in the context of the genetic variability of emerging variants to avoid a potential loss of clinical efficacy.

At the end of December 2021, the World Health Organization (WHO) issued an interim statement on booster doses for COVID-19 vaccination in light of the evidence of waning protection from primary vaccination series (24). Thus, the low nAbs responses against Mu and Delta in all the tested vaccines support the need for a booster dose to improve the protection against SARS-CoV-2 infection or severe COVID-19. However, it remains to be studied in what proportion this will improve the magnitude of nAb titers and clinical efficacy. With the worldwide emergence of SARS-CoV-2 variants with a significant capacity to escape from vaccine-elicited nAbs, a lowered vaccine efficacy is expected. Hence other non-pharmacological measures should be retained in the vaccinated population. The results obtained here, added to the high frequency of the Mu variant during the devastating third pandemic peak registered in Colombia in 2021 (5), highlight the importance of carrying out studies on sera from patients with hybrid immunity (natural and vaccine-elicited) to know the effect of this type of exposure in nAb titers.

## Data Availability

All data produced in the present study are available upon reasonable request to the authors

## Acknowledgments

The authors thank the many researchers at the Dirección de Investigación en Salud Pública – INS involved in sample collection used in this work. We thank Rotary International and Charlie Rut Castro for equipment’s donation for sample screening.

## Author Affiliations

From the group of Genómica de Microorganismos Emergentes, Dirección de Investigación en Salud Pública, (D.A.A-D., M.T.H-S., P.T-R., K.L-D., H.A.R-M., D.P-C., M.G., J.R-G., J.D.C., B.H.D-A., T.C. M.M-R.), and Dirección de Producción (A.M-S., E.J.A-R.) Instituto Nacional de Salud, Bogotá; Fundación Banco Nacional de Sangre Hemolife, Bogotá (A.L.M.).

